# The therapeutic effectiveness of Convalescent plasma therapy on treating COVID-19 patients residing in respiratory care units in Baghdad, Iraq

**DOI:** 10.1101/2020.06.24.20121905

**Authors:** Anwar M. Rasheed, Dhurgham F Fatak, Hashim A. Hashim, Mohammed F Maulood, Khulood K Kabah, Yaqoob A. Almusawi, Ahmed S Abdulamir

## Abstract

**Objectives:** The current COVID-19 pandemic needs unconventional therapies to tackle the resulted high morbidity and mortality. Convalescent plasma is one of the therapeutic approaches that might be of benefit.

**Methods:** Forty nine early-stage critically-ill COVID-19 patients residing in RCU of three hospitals in Baghdad, Iraq were included, 21 received convalescent plasma while 28 did not receive, namely control group. Recovery or death, length of stay in hospital, and improvement in the clinical course of the disease were monitored clinically along with laboratory monitoring through SARS-CoV-2 RNA detection via PCR, and SARS-CoV-2 IgG and IgM serological monitoring.

**Results:** Patients received convalescent plasma showed reduced duration of infection in about 4 days, and showed less death rate, 1/21 versus 8/28 in control group. In, addition, all of the patients received convalescent plasma showed high levels of SARS-CoV-2 IgG and IgM 3 days after plasma transfusion. Plasma from donors with high levels of SARS-CoV-2 IgG and donors with positive SRAS-CoV-2 IgM showed better therapeutic results than other donors.

**Conclusions:** Convalescent plasma therapy is an effective mode of therapy if donors with high level of SARS-Cov2 antibodies are selected and if recipients were at their early stage of critical illness, being no more than 3 days in RCU.

## Introduction

SARS-CoV-2 is a novel virus from the family Coronavirdae that caused deadly pandemic, COVID-19 infection, all over the world starting from December 2019 till the time of writing this article, June 2020. Despite all of the trials for finding a suitable therapy for COVID-19, till now no single or cocktail therapy has proved to cure the infection [1]. Remdesivir has shown partial therapeutic efficacy while other drugs have not proved efficient so far and most of them are still under research [2].

No memory immunity in humans can tackle the spread of SARS-CoV-2 towards the lower respiratory tract [3]. Mortality rate of COVID-19 ranges between 3.5-14% in different parts of the world. Hence, SARS-CoV-2 is 30 times more deadly than influenza virus [4]. Moreover, COVID-19 patients need elaborative hospital intensive care for long duration, up to 5 to 6 weeks. Admission of high numbers of critically-ill COVID-19 patients to hospitals in a short time can undermine any health system [5].

Accordingly, any medicine or therapeutic approach has become desperately needed for decreasing morbidity, mortality and length of hospital stay of COVID-19 patients. One of the therapeutic approaches that were recently highlighted is the convalescent plasma (CP). The CP is an old approach used during the Spanish flu pandemic and proved then to be a good mode of treatment [6]. The CP is taken from recovered COVID-19 patients for the sake to use their anti-SARS-CoV-2 antibodies laden plasma to fight the viral infection in critically-ill patients who their immune system can not eradicate infection efficiently by its own [6,7].

In the current pandemic of COVID-19 with the absence of any cure, CP has become a new target for treating COVID-19 patients [7]. In Iraq, due to 30 years of the political and military unrest, the health system is of limited capacity and can not match high numbers of COVID-19 patients, especially the severe and critically ill cases that occupy RCU beds for several weeks. Therefore, it was mandatory to try whatever approach to treat COVID-19 patients and lower death rate and decrease the length of hospital stay to free as much as possible RCU beds for other patients. In the current study, we tested the effectiveness of the CP therapy on 21 early stage critically ill patients residing in COVID-19 RCU wards in Baghdad, Iraq hospitals. The Aim of the current clinical trial was to assess the safety of the convalescent plasma therapy and to assess the clinical and laboratory improvement after the CP therapy compared to age- and sex-matched control group.

## Patients and methods

The current study started on April 3^rd^, 2020 and lasted till June 1^st^ 2020 and was conducted in Baghdad, Iraq. All of the patients were voluntarily included in the study by a written consent from their relatives. The donors were voluntarily recruited in this study. The current study was started after obtaining the ethical and official approvals from the Baghdad-Alkharkh General Directorate of Health.

### Critically-ill COVID-19 patients and their Inclusion and exclusion criteria

Forty nine critically-ill COVID-19 patients were included in the current study. All of the patients were with pneumonia and residing in RCU; As a result of ABO compatibility and limited plasma, 21 of the patients were randomly chosen to take CP, while other age- and sex-matched 28 patients were under the conventional therapy as control group. The included patients, whether in CP or control group, were of age ≥ 18 y with dyspnea and oxygen saturation less than 90% in resting state. All 50 patients were at their first 3 days in RCU either receiving O2 therapy or on ventilators. We aimed at targeting critically-ill patients, either CP or control groups, at their early stages of admission to RCU before developing full-blown ARDS or respiratory and/or multiple organ failure. All of the patients were residing in infectious diseases wards before being transferred to RCU. The patients were residing in Alkarkh general and Alforat hospitals, while only one patient was in Alyarmug hospital.

The exclusion criteria of the COVID-19 patients were: previous allergic history to plasma or its ingredients such as sodium citrate, and cases with serious general conditions, such as severe organ dysfunction, that are not suitable for transfusion or very late stage of the ARDS where CP has proved to be of low therapeutic benefit [7, 8].

### Inclusion criteria for the donors

The donors of CP were two weeks previously recovered patients from COVID-19. The donors should be younger than 50 years, healthy, non-pregnant females, with no comorbidities, and those who showed moderate COVID-19 infections. The donors’ plasma was tested using SARS-CoV-2 IgG Antibody ELISA. The positive donors for COVID-19 IgG (SARS-CoV-2 IgG index >1) were preliminarily included. Then, only the donors with IgG index equal or more than 1.25 were selected; this ensured getting CP with the highest titers of antibodies. In addition, a rapid immunochromatographic test, COVID-19 IgG/IgM test was used to screen the donors and the recipients for the presence of anti-SARS-CoV-2 IgM antibodies.

### Screening and follow-up of the critically-ill COVID-19 patients

The patients of the current study, CP or control groups, were evaluated by the following:

- SARS-CoV-2 PCR at D2, D3, D5, more than D5: the days after of the first day (or D0) of inclusion to the study for the control group or the day of giving CP for the CP group.
- SARS-CoV-2 IgG and IgM antibodies at D0 and D3, and for the donors at the day of taking CP.
- Continuous medical evaluation of patients’ signs and symptoms or any allergic reaction including fever, skin rash, anaphylactic shock, level of dyspnea, respiratory rate, O2 saturation, need for oxygen, need for ventilators, temperature.

### The end points of the current study

#### The end points for the current study were

First, the safety of the CP therapy within the first 3 hours post-transfusion by mainly monitoring the allergic reaction to CP.

Second, the duration, in days, for the patients to convert to SARS-CoV-2 RNA-negative along with improvement in the signs and symptoms of the critical infection, namely relief of severe dyspnea, no need for ventilators or oxygen therapy, declining in fever if any, declining in respiratory rate to less than 30/min, and increased oxygen saturation to more than 93% at rest, so patients can be discharged from RCU to the infectious disease ward; this duration is named the recovery time from critical illness (RTCI).

Third, the survival or death of the patients.

### PCR for SARS-CoV-2 RNA

The definitive diagnosis for COVID-19 was carried out by using real-time RT-PCR assay (QiaAmp RNA mini blood, Qiagen, Germany for RNA extraction and 2019 nCov KogeneBiotech one step PCR kit R6900TD (South Korea) to detect SARS-Cov2 RNA in nasopharyngeal swabs. The PCR instrument used was rotor-gene 6000 cycler, Qiagen, Germany. The thermal cycling program was conducted according to the manufacturer guidelines.

### Detection and evaluation of anti-SARS-Cov2 antibodies

The SARS-CoV-2 IgG semi-quantitatively and SARS-CoV-2 IgM qualitatively were assessed in the serum of both the donors and recipients by using SARS-CoV-2 IgG Antibody ELISA kit DEIASL019 (Creative Diagnostics, USA) and rapid test COVID-19 IgG/IgM test (Biozek, Netherlands), respectively.

For the estimation of SARS-CoV-2 IgG in donors and recipients, 100 ul of the serum were collected. For each 96 well microtiter plate, blank, negative, and positive controls were included. The procedure of ELSIA was conducted according to the guidelines of kit manufacturer SARS-CoV-2 IgG Antibody ELISA kit DEIASL019 (Creative Diagnostics, USA). At the end, the colorimetric reaction was measured at 450 nm wavelength. The positive results are expressed as IgG index which is calculated by dividing OD of the sample over the OD of the negative control. IgG index more than 1 is considered positive. The higher the IgG index is the higher titer of IgG in the serum. SARS-CoV-2 IgG index >1 is positive, 1-1.25 is weakly positive, 1.25-1.5 is moderately positive, and >1.5 is strongly positive.

For the detection of SARS-CoV-2 IgM antibodies, a qualitative strip test was used giving just negative or positive results.

## Results

It is noteworthy to mention that the included patients in this study, CP and control group, showed, to some extent, a male predilection to more severe COVID-19 infection as 75% of males and 53% of females were on ventilators (P=0.1), while there was no predilection of any parameter studied to certain blood group type (P>0.05).

### The safety of CP therapy

All of the patients, who were given CP, were closely monitored for the first 3 hours after CP for detecting and treating early adverse effect especially allergic reactions. None, except one, of the 21 CP patients developed any allergic reaction. A single case developed mild skin redness and itching lasted for 1 hour after taking CP; intramuscular anti-histamine was injected and terminated the mild allergic reaction.

### Comparison between CP and control group

The CP group was compared to the age- and sex-matched control group in terms of recovery time from critical illness (RTCI), days of infection before inclusion to the study, and the whole duration of infection. It was found that the RTCI in CP group, 4.52±2.3 days, was lower than that in control group, 8.45±1.8 days (P<0.01). Hence, CP accelerated the removal of viral infection and recovery in patients for about 4 days. Accordingly, the whole duration of COVID-19 infection in CP group, 19.3±6.9, was lower than in control group, 23.42±6.4 days (P<0.05). On the other hand, there was no significant difference between CP and control groups in regard to age, sex, temperature at D0, and oxygen saturation at D0 (P>0.05), Table 1.

**Table 1:**
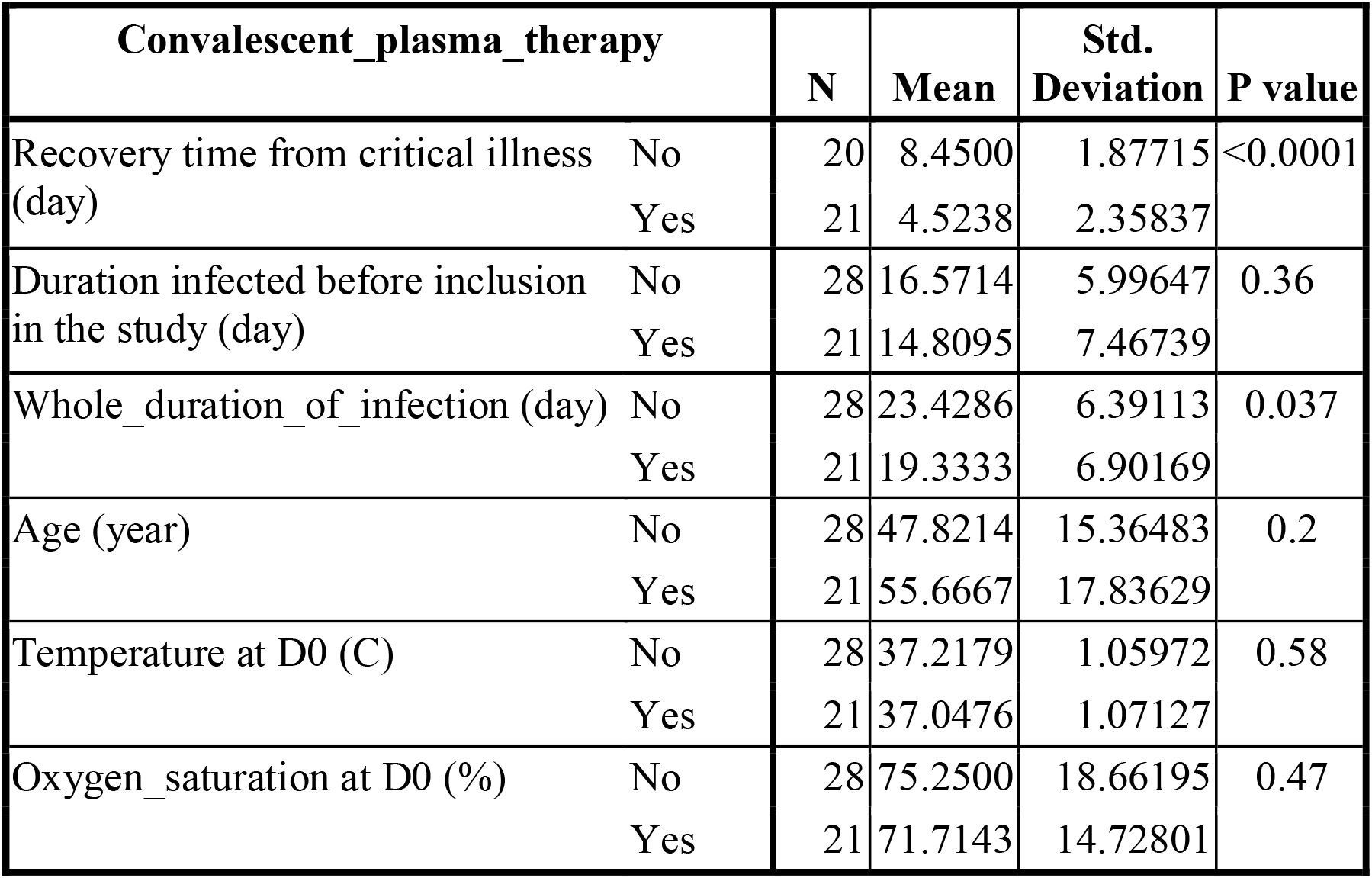
Comparison between Convalescent plasma and control groups in terms of age, recovery time from critical illness, duration of infection before inclusion to the study, the whole duration of infection, presenting temperature and oxygen saturation.

| **Convalescent_plasma_therapy** |  | **N** | **Mean** | **Std. Deviation** | **P value** |
| --- | --- | --- | --- | --- | --- |
| Recovery time from critical illness (day) | No | 20 | 8.4500 | 1.87715 | <0.0001 |
|  | Yes | 21 | 4.5238 | 2.35837 |  |
| Duration infected before inclusion in the study (day) | No | 28 | 16.5714 | 5.99647 | 0.36 |
|  | Yes | 21 | 14.8095 | 7.46739 |  |
| Whole_duration_of_infection (day) | No | 28 | 23.4286 | 6.39113 | 0.037 |
|  | Yes | 21 | 19.3333 | 6.90169 |  |
| Age (year) | No | 28 | 47.8214 | 15.36483 | 0.2 |
|  | Yes | 21 | 55.6667 | 17.83629 |  |
| Temperature at D0 (C) | No | 28 | 37.2179 | 1.05972 | 0.58 |
|  | Yes | 21 | 37.0476 | 1.07127 |  |
| Oxygen_saturation at D0 (%) | No | 28 | 75.2500 | 18.66195 | 0.47 |
|  | Yes | 21 | 71.7143 | 14.72801 |  |

There was no significant difference in the percentage of patients on ventilators, 81% in CP versus 57% in control groups (P>0.05). The level of SARS-CoV-2 IgG at D0 in CP and control groups were not different (P>0.05), while at D3, CP group showed much higher number of patients with positive and strongly positive IgG than in control group (P<0.05), Table 2 and figure1. For IgM at D0, both groups showed very similar percentage of positive SARS-CoV-2 IgM, 19% in CP versus 18% in control groups, (P>0.05). For IgM at D3, CP group showed much higher number of patients with positive IgM than in control group (P<0.05), Table 3, figure 2. Most importantly, the death rate in CP group was much lower than in control group. Only one 1/21 (4.8%) patient in CP group died versus 8/28 (28.5%) in control group (P<0.05), table 4, figure 3.

**Table 2:**
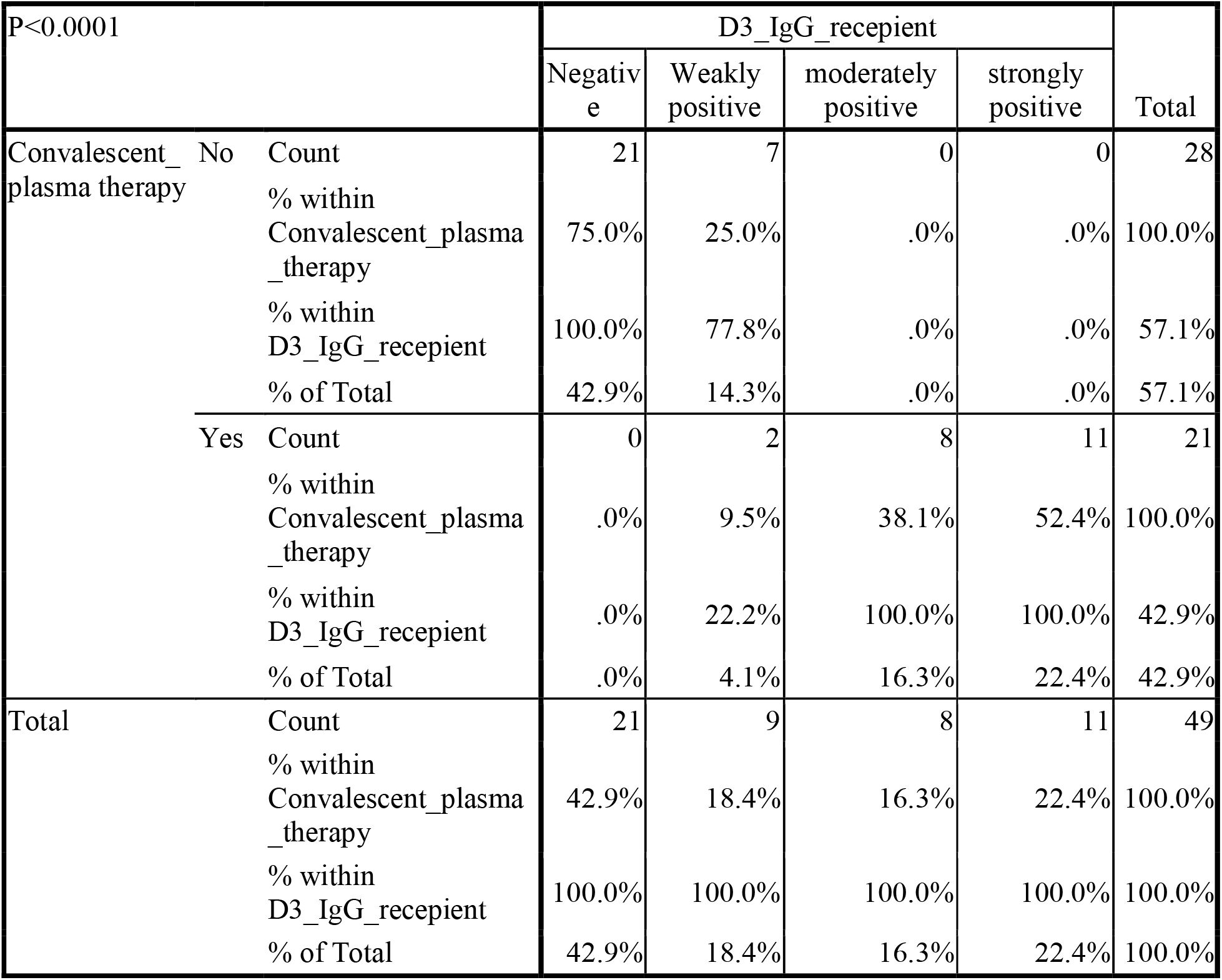
Number (%) of seropositivity level of SARS-CoV-2 IgG 3 days after inclusion in the study for control group or 3 days after taking plasma in Convalescent plasma group

**Table 3:** Number (%) of seropositivity level of SARS-CoV-2 IgM 3 days after inclusion in the study for control group or 3 days after taking plasma in Convalescent plasma group

| P<0.0001 |  |  | D3_IgM_recepient |  | Total |
| --- | --- | --- | --- | --- | --- |
|  |  |  | Negaivet | Positive |  |
| Convalescent_plasma<br>_therapy | No | Count | 20 | 8 | 28 |
|  |  | % within Convalescent_plasma_t<br>herapy | 71.4% | 28.6% | 100.0% |
|  |  | % within D3_IgM_recepient | 100.0% | 27.6% | 57.1% |
|  |  | % of Total | 40.8% | 16.3% | 57.1% |
|  | Yes | Count | 0 | 21 | 21 |
|  |  | % within Convalescent_plasma_t<br>herapy | .0% | 100.0% | 100.0% |
|  |  | % within D3_IgM_recepient | .0% | 72.4% | 42.9% |
|  |  | % of Total | .0% | 42.9% | 42.9% |
| Total |  | Count | 20 | 29 | 49 |
|  |  | % within Convalescent_plasma_t<br>herapy | 40.8% | 59.2% | 100.0% |
|  |  | % within D3_IgM_recepient | 100.0% | 100.0% | 100.0% |
|  |  | % of Total | 40.8% | 59.2% | 100.0% |

**Table 4:**
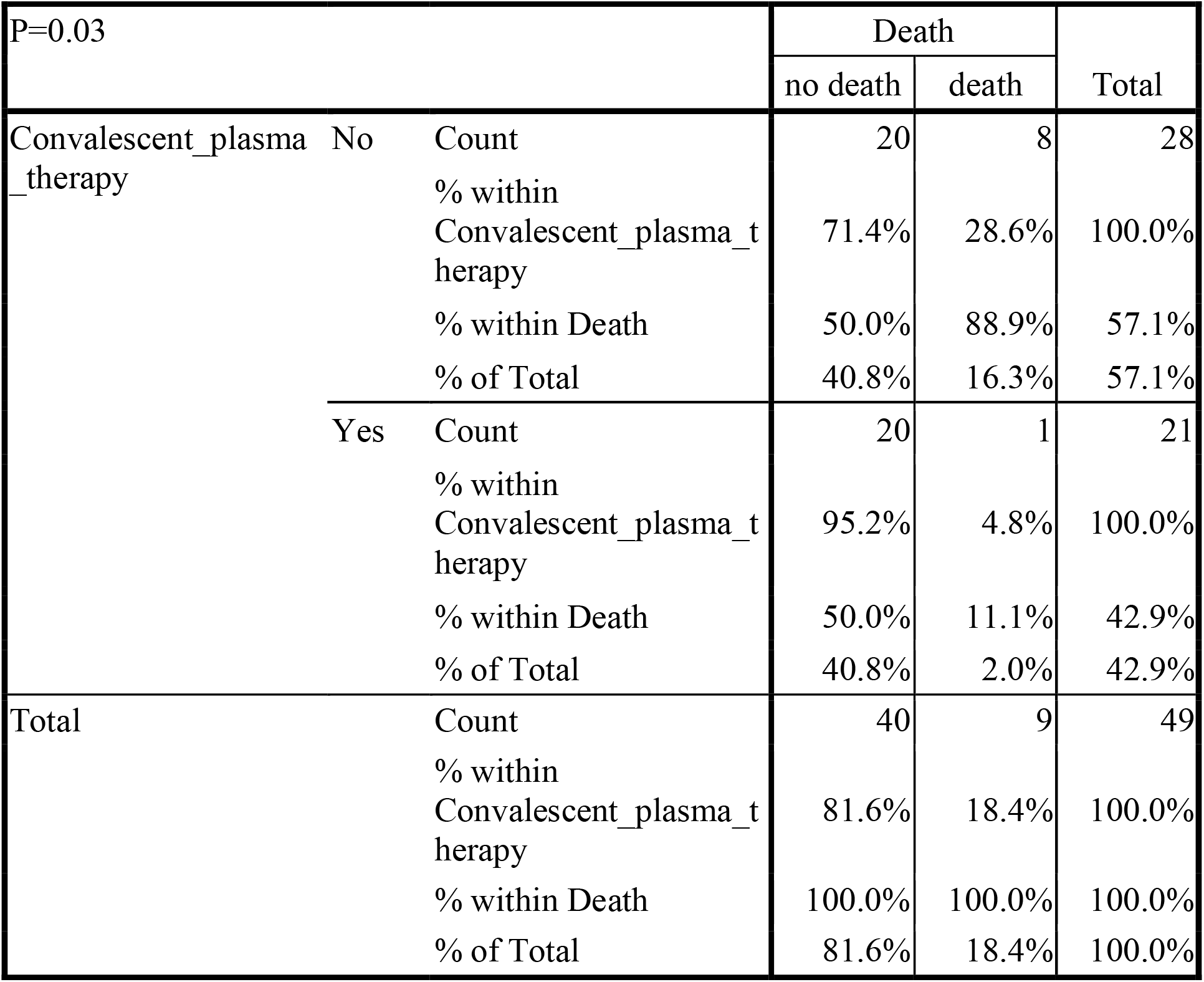
Death rate in Convalscent plasma group versus control group.

**Figure 1:**
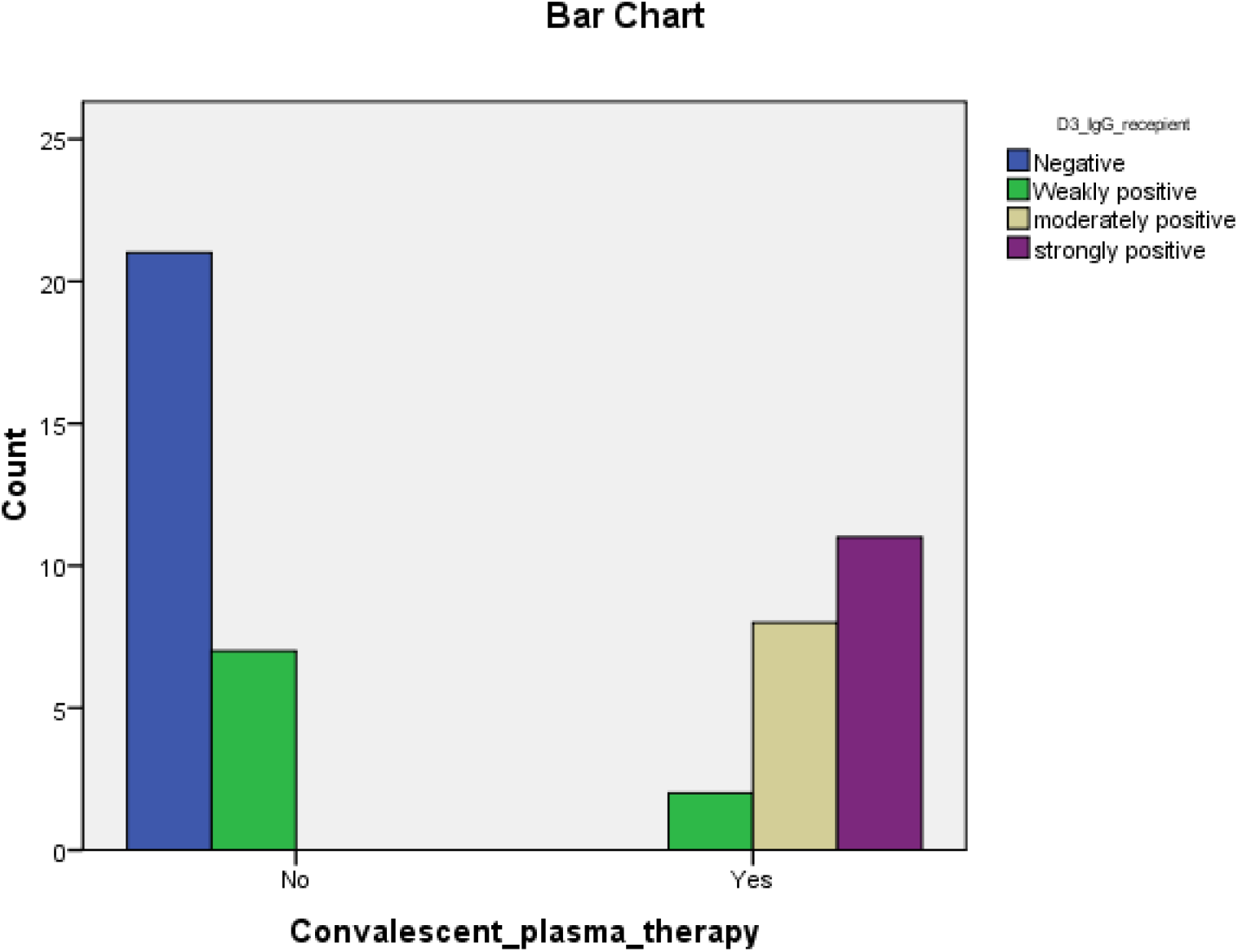
Number of patients with negative, weakly positive, moderately positive, and strongly positive serum SARS-CoV-2 IgG after 3 days from inclusion in the study between Convalescent plasma versus control groups.

**Figure 2:**
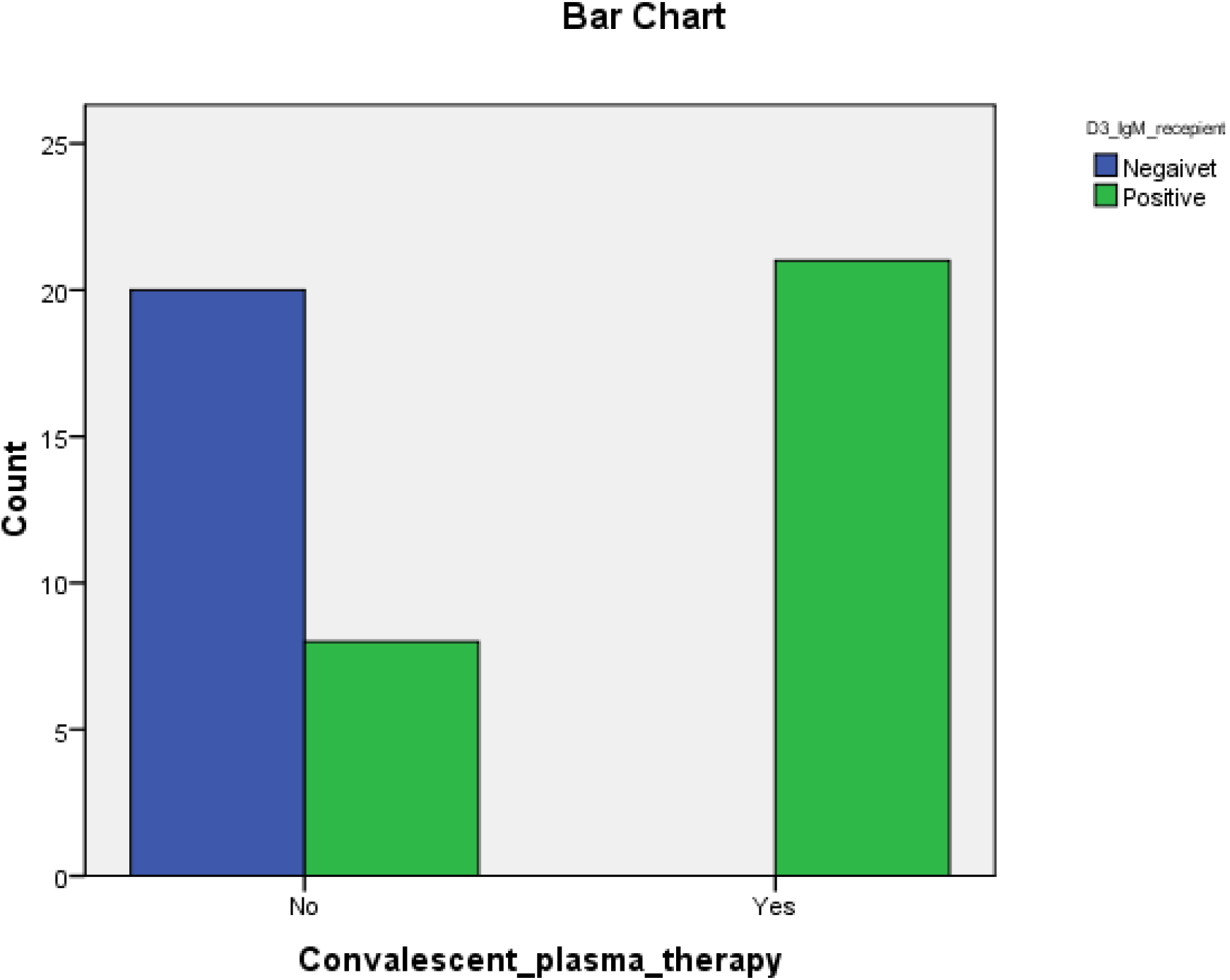
Number of patients with negative and positive serum SARS-CoV-2 IgM after 3 days from inclusion in the study between Convalescent plasma versus control groups.

**Figure 3:**
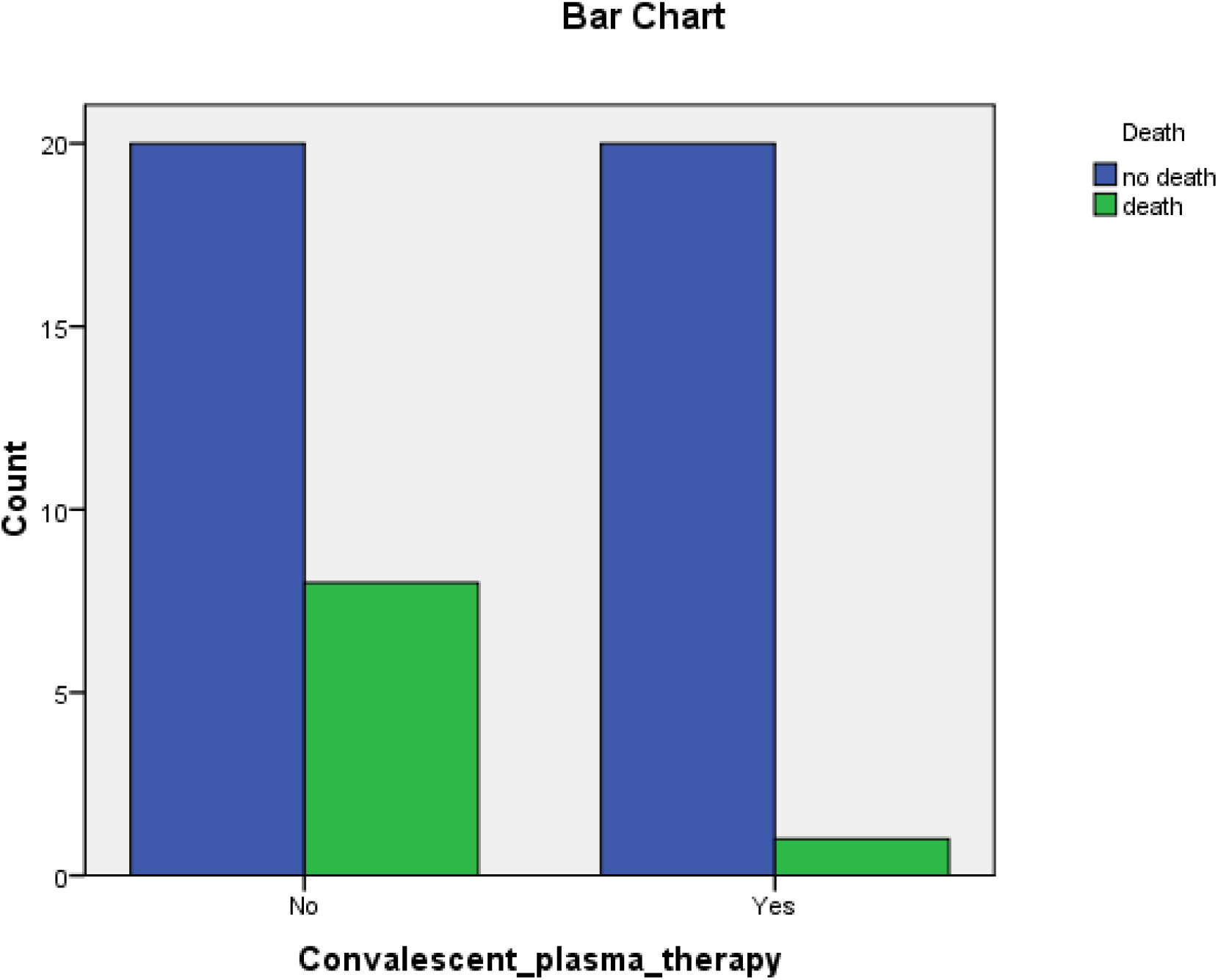
The count of death versus no death in Convalescent plasma and control groups.

### The studied parameters in CP group and the effect of donors’ plasma

The parameters studied in the CP group are listed in table 5. The CP group consisted of 57.1% males and 42.86% females. Blood groups of CP patients and control patients were B+ 47.6%, O+ 23.8%, and each of A+ and O-14.3%. 81% of them on ventilator while the rest on oxygen therapy. All of the CP patients were early admitted from infectious diseases ward to RCU (not more than 3 days).

**Table 5:**
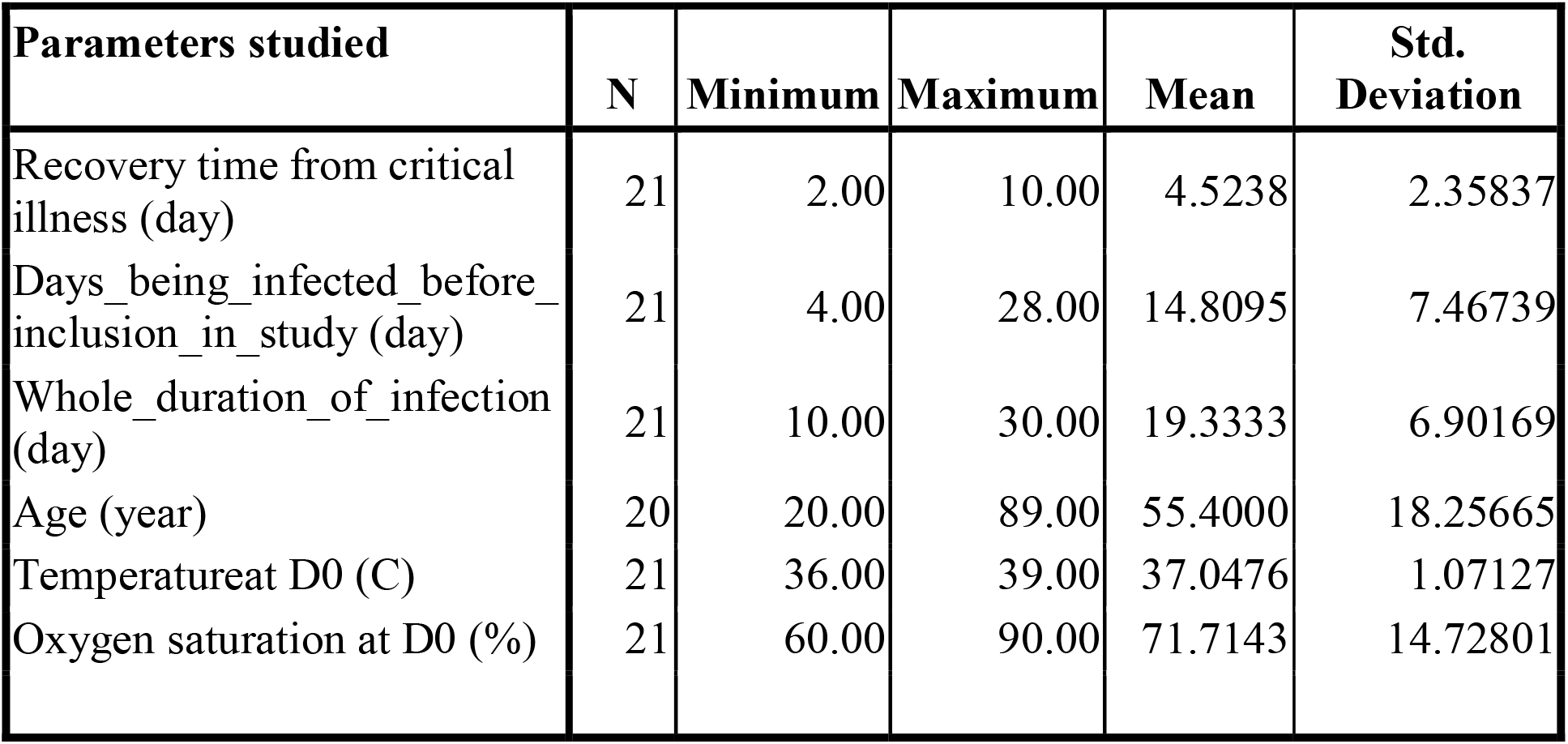
The studied parameters in the Convalescent plasma group of patients.

| **Parameters studied** | **N** | **Minimum** | **Maximum** | **Mean** | **Std.**<br>**Deviation** |
| --- | --- | --- | --- | --- | --- |
| Recovery time from critical illness (day) | 21 | 2.00 | 10.00 | 4.5238 | 2.35837 |
| Days_being_infected_before_inclusion_in_study (day) | 21 | 4.00 | 28.00 | 14.8095 | 7.46739 |
| Whole_duration_of_infection (day) | 21 | 10.00 | 30.00 | 19.3333 | 6.90169 |
| Age (year) | 20 | 20.00 | 89.00 | 55.4000 | 18.25665 |
| Temperature at D0 (C) | 21 | 36.00 | 39.00 | 37.0476 | 1.07127 |
| Oxygen saturation at D0 (%) | 21 | 60.00 | 90.00 | 71.7143 | 14.72801 |

At the day of taking CP or D0, only 14.9% were weakly positive to SARS-CoV-2 IgG while the rest were negative; on the other hand, at D3, zero patients were negative to SARS-Cov2 IgG, 9.52% weakly positive, 38.1% moderately positive, and 52.38% strongly positive (P<0.05),. For IgM, only 19% were positive at D0 versus 100% positive at D3 (P<0.05).

For the donors of CP, depending on the availability of donors, we tried as could as possible to select the highly positive plasmas for SARS-CoV-2 IgG. Up to 52% of donors were moderately positive and 48% of them were strongly positive for SARS-CoV-2 IgG. In addition, up to 53% of the donors were shown to be positive to SARS-CoV-2 IgM.

Actually, only one case died in CP group; and this case had nothing peculiar in regard to the presenting temperature or the whole duration of infection (P>0.05). This patient died after 4 days of taking CP. However, this case was at the end stage of the disease with oxygen saturation at 60% and with severe comorbidity and immunosuppression due to cancer and chemotherapy.

The SARS-CoV-2 IgG at D0 in recipients was shown to be associated with the RTCI (P<0.05). The mean RTCI of positive SARS-CoV-2 IgG recipients at D0, was only 2.3±0.58 days, while the mean RTCI at D0 of negative SARS-CoV-2 IgG patients was 4.8±1.3 days after taking CP, figure 4. In addition, the positive IgG CP patients at D0 were those who were infected for longer time before taking CP, mean 22.6±7.9 days compared to the D0 IgG negative, 13.5±6.5 days (P<0.05). Hence, the patients who took longer time of infection before being deteriorated and given CP developed some level of SARS-CoV-2 IgG at time of taking CP and were shown to be more responsive to CP therapy.

**Figure 4:**
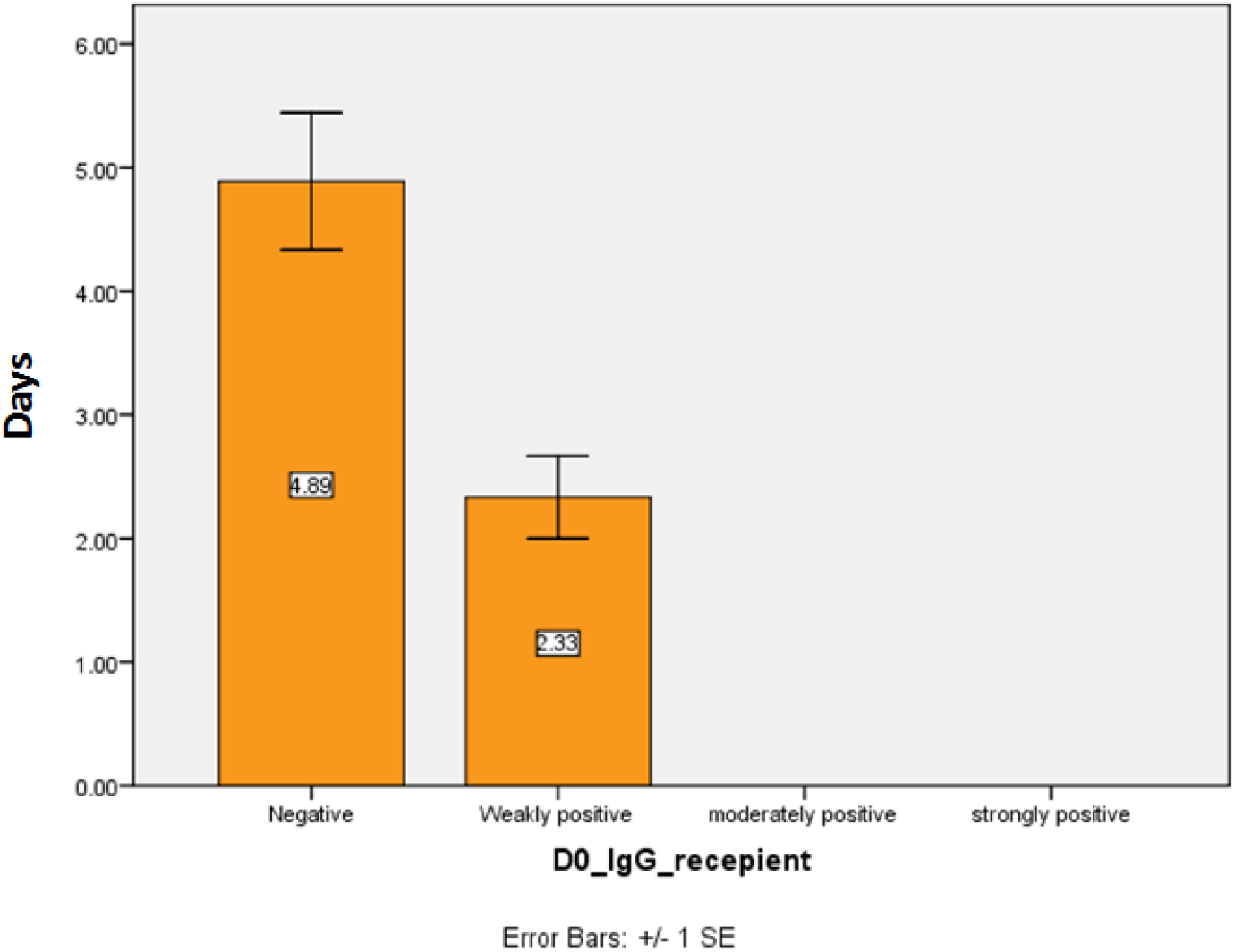
The mean Recovery time from critical illness, in days, in Convalescent plasma recipients with negative versus weakly positive SARS-CoV-2 IgG at D0

Similarly, CP patients who developed stronger SARS-CoV-2 IgG at D3 were more prone to benefit from CP therapy. The mean RTCI were 6.3±2.3, 4±0.01, and 3.2±1.6 days in CP patients who developed weakly positive, moderately positive, and strongly positive SARS-CoV-2 IgG at D3, respectively (P=0.009), figure 5. On the other hand, SARS-CoV-2 IgM at D0 or D3 in recipients of CP did not show any effect on the outcome of CP therapy (P>0.05).

**Figure 5:**
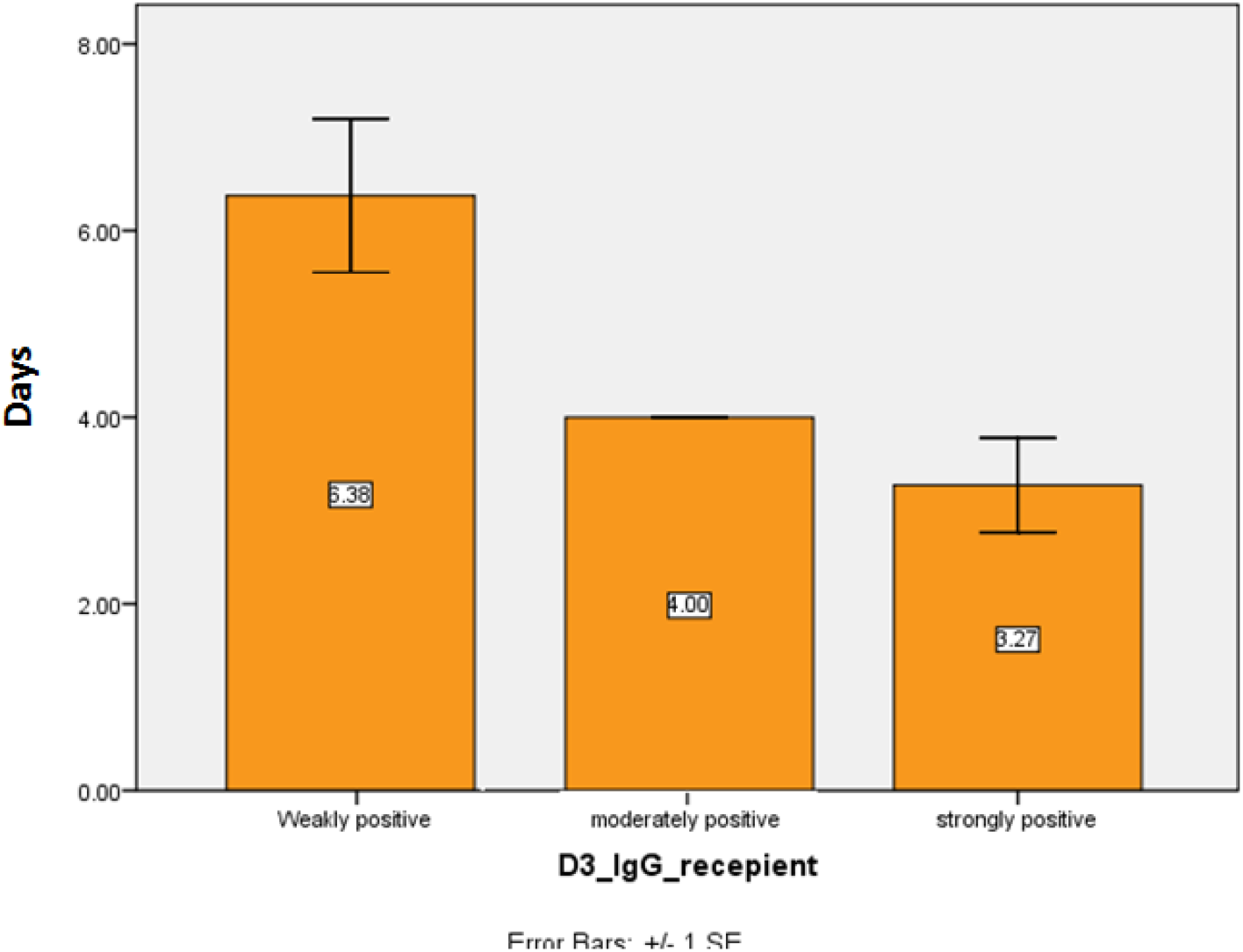
The mean Recovery time from critical illness, in days, among Convalescent plasma recipients with weakly, moderately, and strongly positive SARS-CoV-2 IgG at D3.

Interestingly, the donor plasma’ level of SARS-CoV-2 IgG and the presence of SARS-CoV-2 IgM were found to be of effect on the recovery of COVID-19 treated with CP. The mean RTCI was much lower, 3.18±1.4 days, in patients received CP from SARS-CoV-2 IgM positive donors, than in recipients took CP from SARS-CoV-2 IgM negative donors, 6±2.3 days (P=0.003), figure 6. Alike, the mean RTCI was lower, 3.6±1.9 days, in patients received CP from donors with strongly positive SARS-CoV-2 IgG, than in recipients received CP from donors with moderately positive SARS-CoV-2 IgG, 5.36±2.2 days (P=0.048), figure 7. Hence, the titer of SARS-CoV-2 IgG and the presence of SRAS-CoV-2 IgM antibodies in the convalescent plasma of donors seem to play important role for the success of CP therapy.

**Figure 6:**
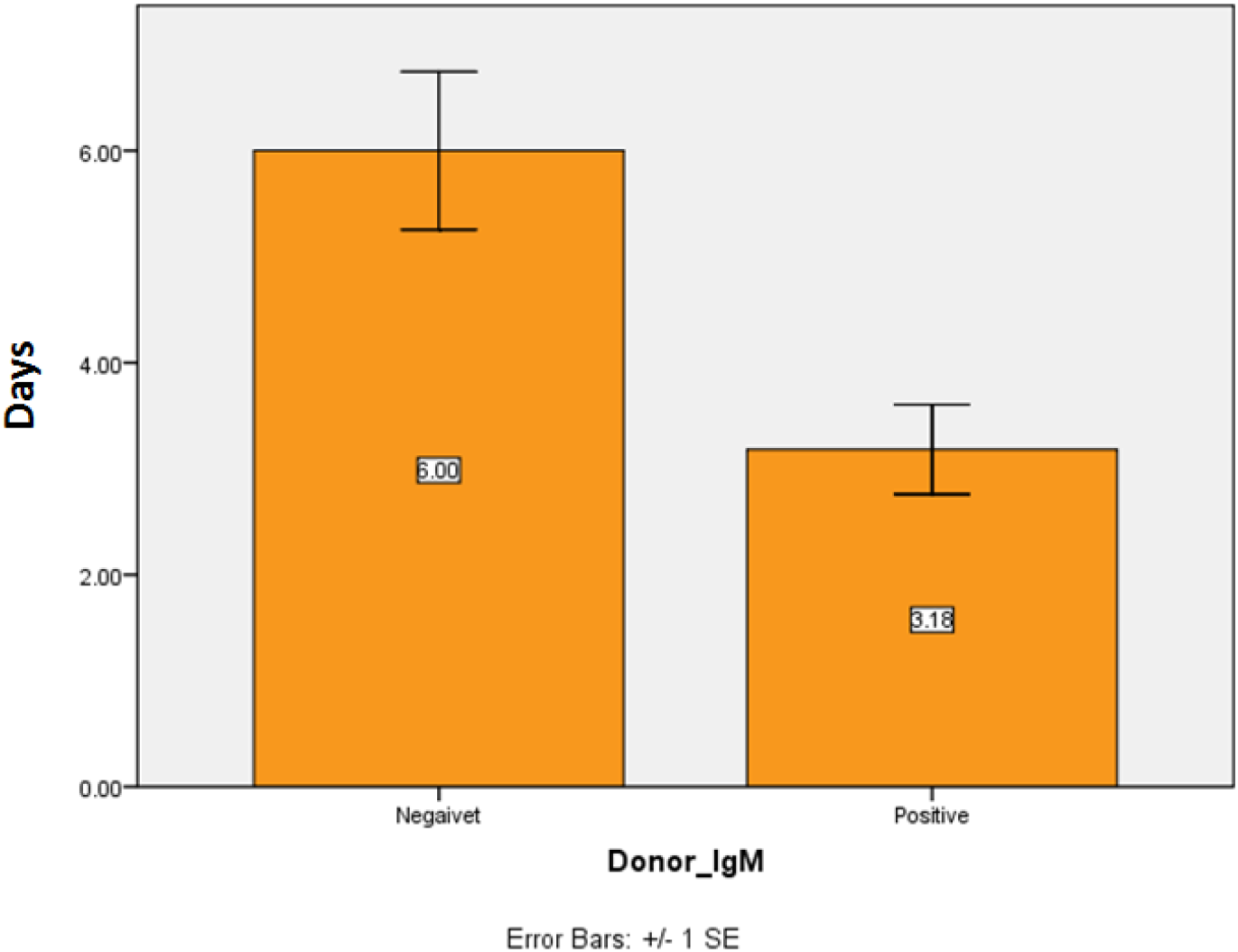
The mean Recovery time from critical illness, in days, in recipients of Convalescent plasma from donors negative versus positive SARS-CoV-2 IgM

**Figure 7:**
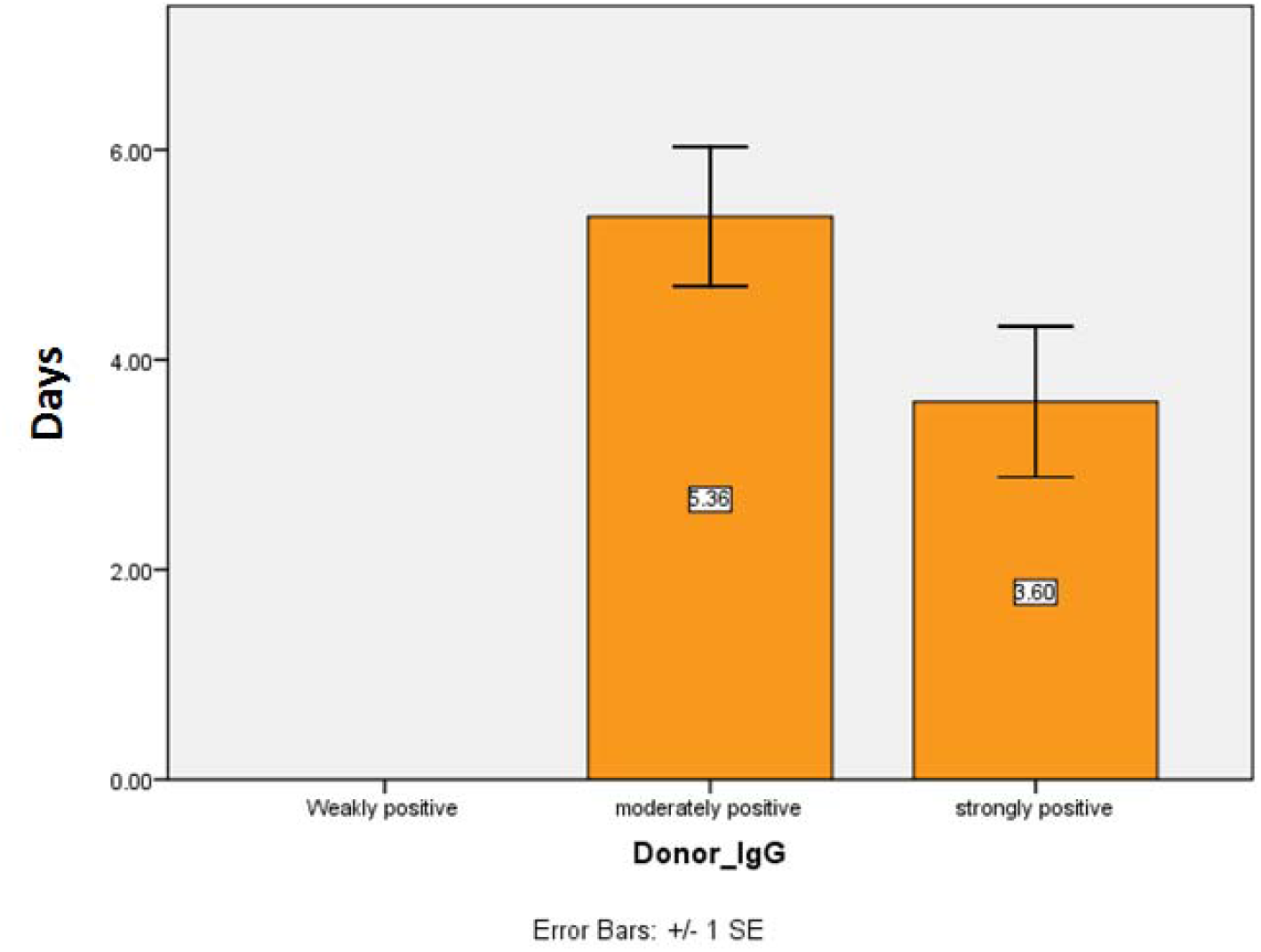
The mean Recovery time from critical illness, in days, in recipients of Convalescent plasma from donors moderately positive versus strongly positive SARS-CoV-2 IgG

## Discussion

CP was shown to be safe in the current study and lowered death rate from 28% in control group to only 4.8% in CP group. This is a remarkable lowering in mortality rate, which is about 23% less in patients received CP. In the current study, the observed effect of CP in reducing mortality rate might be attributed to two main factors. First, the target patients of this study were patients who were recently deteriorated and admitted to RCU, no more than 3 days in RCU. It is thought that if CP is given early to critically-ill patients, the effect of CP in treating the infection and preventing further lower respiratory tract damage would be noticeable. A previous study found that CP given to 6 critically-ill COVID-19 patients managed to eradicate virus 100% in 2-3 days after taking plasma, but only 1/6 of the patients was saved [7]. This was due to the fact that those 6 patients were given CP at advanced stage of the disease when ARDS became full-blown [7-9]. Therefore, the current study was designed to avoid targeting very advanced COVID-19 patients. The second possible factor for the good outcome observed in the current study, the level of SARS-CoV-2 IgG and IgM in donors was monitored and only the plasma with highest levels of SARS-CoV-2 IgG were selected; in addition, only young and quite healthy donors were chosen.

In addition to reducing the death rate, CP in the current study decreased the time needed for the critically-ill COVID-19 patients to recover. CP in the current study accelerated the recovery of patients in about 4 days. In fact, this is an important outcome as freeing RCU beds to other critically-ill patients is vital during COVID-19 pandemic.

It is noteworthy to mention that the positive SARS-CoV-2 IgG recipients of CP at D0, benefited more from the immune boosting effect of CP therapy than those who were negative at D0. This was shown by the less days needed to recover and convert to SARS-CoV-2 RNA negative in the recipients who were already positive in IgG antibodies. This actually can be one of the predictors for the success of CP therapy. Furthermore, the serum level of SARS-CoV-2 IgG 3 days after taking CP was shown to have a direct effect on the time needed to recover after CP. All these measured parameters highlights that CP is an efficient boosting therapy for the immune system of critically-ill patients and it is a useful mean to elevate anti-SARS-CoV-2 antibodies with enhanced rates of recovery and survival.

In addition, the current study indicated that the presence of SARS-CoV-2 IgM in the plasma of donors reduced the time for recovery from 6 days to 3.6 days after taking CP. And for IgG in the donors, the strongly positive donors helped the recipients to recover earlier, 3.6 days instead of 5.3 days, after taking CP. Therefore, keen selection of donors for CP is vital for maximizing the therapeutic effect of this approach. Several studies reported that the titer of SARS-CoV-2 IgG in the plasma of donors is important in the outcome of CP therapy [7-11].

Taken together, CP therapy proved to be a good therapeutic approach if the plasma of donors were rich with SARS-CoV-2 IgG and IgM. In addition, the recipients who have already started producing SARS-IgG and IgM are more prone to benefit from CP. Furthermore, maybe the most important factor, targeting critically-ill patients at early days in RCU and on ventilators seems vital for the success of CP therapy. Too late CP therapy might be good to eradicate the virus, if any, but it might not be enough to reverse the advanced inflammatory damage in the lung and/or other organs in COVID-19 patients. CP therapy proved in this study to lower mortality and morbidity and to accelerate recovery. Therefore, the observed outcome of CP therapy is encouraging to be trialed on higher number of patients.

## Data Availability

The data are available

## Acknowledgment

This study was financed, approved and supported by the Karkh health directorate in Bghdad, Iraq and was conducted in three main hospitals, Alkarkh hospital, Alforat Hospital and Alyarmug Hospital. This study owes much to the patients and all of the medical and paramedical staff working in the above mentioned hospitals.

